# Transmission of SARS-CoV-2 by children to contacts in schools and households: a prospective cohort and environmental sampling study in London

**DOI:** 10.1101/2021.03.08.21252839

**Authors:** Rebecca Cordery, Lucy Reeves, Jie Zhou, Aileen Rowan, Patricia Watber, Carolina Rosadas, Michael Crone, Marko Storch, Paul Freemont, Lucy Mosscrop, Alice Cowley, Gina Zelent, Kate Bisset, Holly Le Blond, Sadie Regmi, Christian Buckingham, Ramlah Junaideen, Nadia Abdulla, Joseph Eliahoo, Miranda Mindlin, Theresa Lamagni, Wendy Barclay, Graham P. Taylor, Shiranee Sriskandan

## Abstract

**Background:** Assessing transmission of SARS-CoV-2 by children in schools is of critical importance to inform public health action. We assessed frequency of acquisition of SARS-CoV-2 by contacts of children with COVID-19 in schools and households, as well as the amount of virus shed into the air and onto fomites in both settings.

**Methods:** Cases of COVID-19 in children in London schools were identified via notification. Weekly sampling for 3-4 weeks and PCR testing for SARS-CoV-2 of immediate classroom contacts (the “bubble”), non-bubble school contacts, and household contacts was undertaken supported by genome sequencing, along with surface and air sampling in the school and home environment.

**Results:** Within schools, secondary transmission was not detected in 28 individual bubble contacts, representing 10 distinct bubble classes. Across 8 non-bubble classes, 3/62 pupils tested positive– all three were asymptomatic and tested positive in one setting on the same day, unrelated to the original index case. In contrast, the secondary attack rate in naïve household contacts was 14.3% (5/35) rising to 19.1% (9/47) when considering all household contacts. Environmental contamination with SARS-CoV-2 was rare in schools, regardless of school type; fomite SARS-CoV-2 RNA was identified in 4/189 (2.1%) samples in bubble classrooms, 2/127 (1.6%) samples in non-bubble classrooms, and 5/130 (3.8%) samples in washrooms. This contrasted with fomites in households, where SARS-CoV-2 RNA was identified in 60/248 (24.2%) bedroom samples, 66/241 (27.4%) communal room samples, and 21/188 (11.2%) bathroom samples. Air sampling identified SARS-CoV-2 RNA in just 1/68 (1.5%) of school air samples, compared with 21/85 (24.7%) of air samples taken in homes.

**Summary:** The low levels of environmental contamination in schools are consistent with low transmission frequency and adequate levels of cleaning and ventilation in schools during the period of study. Secondary transmission in schools was rare. The high frequency of secondary transmission in households associated with evident viral shedding throughout the home suggests a need to improve advice to households with infection in children in order to prevent onward community spread by sibling and adult contacts. The data highlight that transmission from children is very likely to occur when precautions are reduced.

## Introduction

The potential for schools to amplify outbreaks is well-recognised (1-3). School closures were associated with a reduction in COVID-19 incidence and mortality at the start of the pandemic (4,5) albeit effects remain confounded by other non-pharmaceutical interventions. Importantly, any benefits of closures must be weighed against the unquestionable harms to children and wider society.

COVID-19 poses a much lower risk to children than to adults, both in terms of illness severity (6) and risk of acquisition; children appear half as likely as adults to acquire SARS-CoV-2 (7). The onward transmission risk from SARS-CoV-2-infected children has been subject to less rigorous evaluation, though shedding of virus by children is not markedly different to adults (8). Point prevalence studies indicating a low prevalence of SARS-CoV-2 in schools (9) makes large scale monitoring of transmission impractical and uneconomic. Although outbreaks provide an opportunity to study onward transmission, these are often complicated by uncertainty about timing and transmission direction; one study in schools did identify children as a source of onward transmission in a small number of cases, but such outbreaks comprised just two cases on average (10).

Forward contact tracing offers an opportunity to search actively for secondary infections in a controlled manner. Despite this, transmission of respiratory infection in schools is seldom quantified except in the context of major outbreaks. Clinical attack rates of 20-30% are reported in schools affected by influenza A (11), however the role of silent infection and onward transmission from such cases is not well-studied. In a scarlet fever contact tracing study, we found that outbreak strains spread to over one-quarter of classroom contacts, despite treatment and isolation of index cases. (12) The potential for classrooms and asymptomatic ‘shedders’ to act as an accelerator for respiratory infection is therefore undeniable.

We adapted our contact-tracing protocol to investigate transmission of SARS-COV-2 by children in schools and households. The TraCK (<u>Tra</u>nsmission of <u>C</u>oronavirus-19 in <u>K</u>ids, ISRCTN 13773960) study aimed to assess the risk posed by a SARS-CoV2-infected child who attends school, via longitudinal sampling of the child, school and household contacts, and associated environments, to evaluate and inform interventions to limit spread of COVID-19.

## Methods

### Study eligibility

Schools in London reporting new cases of SARS-CoV-2 infection to local Health Protection teams were invited to take part if a child (index case) had been attending school in the 48h prior to a positive PCR test for SARS-CoV-2. Contextual information relating to prevailing regulations are in Appendix p8. Parents/guardians of notified cases were invited to allow their child and wider household to participate in the study. If the school was willing to support the study, parents/guardians of contacts were also invited to allow their child to participate in the study. The study commenced October 9^th^ 2020 and recruitment ended July 18^th^ 2021.

### Case definition

Children aged 2-14 years (extended to <18 years in November 2020) with a new nose and/or throat swab PCR test result positive for SARS-CoV-2 from an accredited laboratory. Findings from cases will be reported elsewhere.

### Contact definition

Bubble contacts (BC) were children identified by schools who were required to isolate at home due to direct contact with a case. Non-bubble school contacts (SC) were children from a different ‘control’ class in the same school. Household contacts (HC) were adults and children of any age normally resident with the case, and required to isolate.

### Contact sampling

Combined nose and throat samples (single swab of throat followed by nostrils) were taken by the research team from each participating contact (BC, SC, or HC) as soon as possible (<48 hours) after case identification, and thereafter weekly for a total of 4 visits (3 visits from December 2020).

### Environmental sampling

In households, surface and air samples were obtained in each of three rooms (child’s bedroom, communal room, bathroom) at the first visit and thereafter weekly for a minimum of 4 visits (3 visits from December 2020); in some households sampling was undertaken more frequently in the first two weeks. In schools, surface and air samples were obtained weekly from the bubble classroom, school contact classroom, and washrooms. (for details see Appendix p9-10)

### Virological testing

Nasopharyngeal swabs were tested for SARS-CoV-2 E-gene RNA and human RNAseP RNA by an accredited, quantitative RT-PCR followed by genome sequencing (Appendix p9-10). (13). Results were reported in real-time to participants and positive results subject to statutory reporting and associated regulations. Environmental samples were tested by a research laboratory (14) (Appendix p10).

### Gingival crevicular fluid (GCF)

GCF samples were collected from contacts on each sampling occasion (Appendix p11) then tested for total IgG against SARS-CoV2 nucleoprotein by the reference laboratory (15).

### Ethical approval

The study was approved by a research ethics committee (Schools Transmission Study REC reference 18/LO/0025; IRAS Reference 225006). Written, informed consent was obtained from all participants or parents/guardians, and assent was obtained from participants aged under 18.

### Statistical analysis

Analysis was mostly descriptive due to sample size (Appendix p9); Fisher’s exact test was used to compare proportions of household contacts with positive results (Stata version 15). Human target RNAs were compared using Mann Whitney U test (GraphPad Prism).

## Results

Eight schools participated, of which 5 were primary, 2 secondary, and 1 was a special educational needs (SEND) school. In the course of the study, 428 combined nose and throat swabs and GCF samples were obtained from contacts of index cases. Environmental sampling included a total of 1620 surface samples, of which 446 were from schools, and 218 air samples, of which 68 were from schools.

### Transmission to Bubble Contacts

BC were recruited from 10 bubbles in 8 schools. In total 28 bubble contacts who were required to quarantine at home, were followed weekly. Onward transmission of SARS-CoV-2 to the 28 participating BC was not detected over the sampling period (Figure 1A, Table 1). Only 4/28 (14.3%) BC had evidence of prior exposure to SARS-CoV-2 from GCF testing. In one setting, a non-participating BC developed a fever and reported a positive community test. That child was recruited as a ‘case’ along with their household. Subsequent study sample PCR tests were negative, but GCF seroconversion at 4 weeks was consistent with this child being a co-primary case in the class. Participation rate among BC in each school varied widely (median 8.5%, range 2.4% - 26.9%), being lowest in SEND and secondary schools.

**Table 1.** Transmission to Bubble classroom contacts.

| School | Bubble size (incl. cases) | Case number Bubble exposed to | Bubble participant number | Number of bubble contacts testing PCR positive<sup>†</sup> |  |  |  | Bubble contact crevicular fluid anti-NP total IgG |  |
| --- | --- | --- | --- | --- | --- | --- | --- | --- | --- |
|  |  |  |  | Week 1 | Week 2 | Week 3 | Week 4 | Positive on first sample | Positive on last sample |
| A | 29 | 1 | 3 | 0/2 | 0/3 | 0/3 | 0/3 | 1/3 | 1/3 |
| B | 26 | 1 | 7 | 0/4 | 0/4 | 0/6 | 0/7 | 2/7 | 1/7 |
| D | 41 | 1 | 1 | 0/1 | 0/1 | ND | 0/1 | 0/1 | 0/1 |
| E<sup>‡</sup> | 39 | 1 | 2 | 0/2 | 0/2 | 0/2 | ND | 0/2 | 0/2 |
| F<sup>§</sup> | 48 | 2 | 5 | 0/5 | 0/5 | 0/5 | ND | 0/5 | 0/5 |
| G<sup>¶</sup> | 16 | 4 | 2 | 0/2 | 0/2 | 0/2 | 0/2 | 0/2 | 0/2 |
| K<sup>†</sup> | 150 | 11 | 6 | 0/6 | 0/6 | 0/6 | ND | 1/6 | 1/6 |
| M | 30 | 1 | 2 | 0/1 | 0/2 | 0/2 | ND | 0/2 | 0/2 |
| TOTAL |  |  | 28 | 0/23 | 0/25 | 0/26 | 0/13 | 4/28 | 3/28 |

<sup>‡</sup>swabbing delayed until 7d after case confirmed.
<sup>§</sup>Includes 2 different bubbles exposed to one case each. One non-participant bubble contact tested positive in community test (included in household study).
<sup>¶</sup>Bubble exposed to 2 adult and 2 child cases
<sup>†</sup> Includes 2 different bubbles exposed to 4 cases and 7 cases
ND, not done

**Figure 1.**
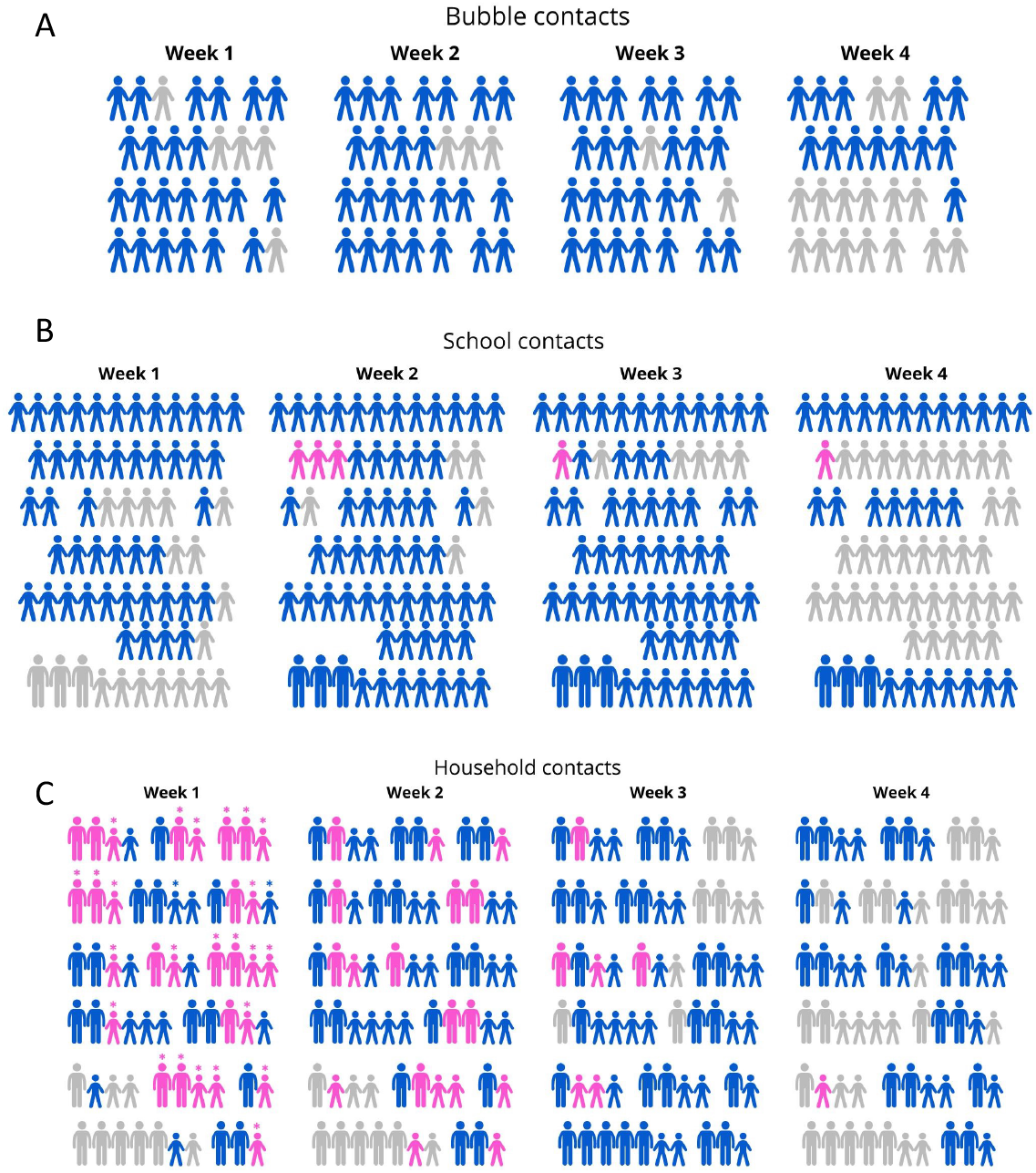
Pictograms of individual contacts in each week of sampling. A. Bubble contacts (n=28). B. School contacts (n=62 pupils, 3 staff) and C. Household cases and contacts. (n=63). For panel C, the 26 participants reported to the study team as having tested positive prior to research swabbing are indicated by* (child index cases, adult and child household contacts). Colour of icons indicates research swab test result in each week of study: Blue icons,negative swab result; pink icons, SARS-CoV2 detected; grey icons, subject not swabbed in that week or not recruited yet. Two of three pupils identified incidentally are included in both panels B and C (i.e. school and household contact pictograms); although pupils were swabbed weekly, the associated households were recruited only after week 2. Within each panel, the figure position is consistent in each week and represents individual participants so can be compared between weeks 1-4. Individual settings are separated by gaps between groups of figures. For presentation purposes, the ordering of settings between panels A, B and C is not the same. Longitudinal sampling was limited to three weeks rather than four weeks for part of the study hence some subjects were not swabbed in week 4.

### Transmission to and between non-bubble School Contacts

Sixty-two pupil SC and 3 staff were recruited from the same 8 schools. SC participation rates were higher than BC, median 22.4% (range 5.2-54.5%). Of those tested, 13/65 (20%) had GCF antibodies indicating previous SARS-CoV-2 infection.

In 7/8 participating schools, no SC were found to be infected with SARS-CoV-2. In setting E, a secondary school, all SC tested negative in week 1, but in week 2, unexpectedly, SARS-CoV-2 was detected in swabs of 3/10 SC. (Figure 1B, Table 2). All three were asymptomatic; in one, the viral load increased from 293,240 E gene copies/swab to 5,999,560/swab copies 3 days later and onward transmission to a sibling household contact who shared a bedroom (84,040 E gene copies/swab) was observed. The other two asymptomatic SC had very low viral loads; the first had 280 E gene copies/swab but was tested only once. The second had 560 E gene copies/swab; samples 7 days earlier, and 4 days later were PCR-negative; and anti-SARS-CoV-2 antibodies were already present in GCF in weeks 1 and 2. It was felt possible that these low viral levels did not represent true infections, but transient mucosal contamination while in the company of a fellow pupil with active infection. The original index case in setting E had been identified following a community PCR test; by week 1 of SC testing the index case had a negative PCR test and was still quarantined. It was inferred that the infection in SC was not linked directly to the original index case.

**Table 2.**
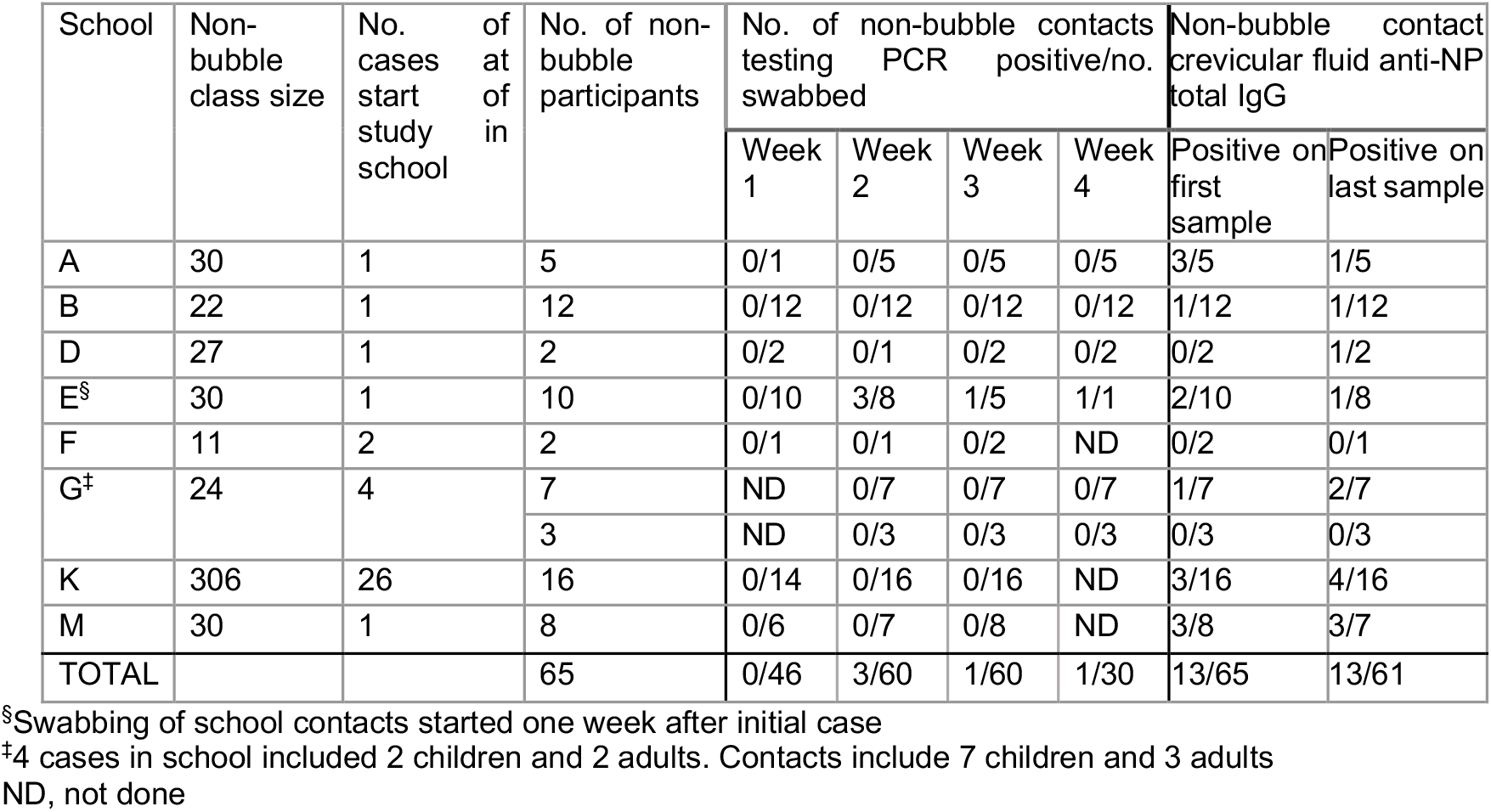
Transmission to non-bubble (control) class contacts.

### Transmission to Household contacts

Sixteen households took part, comprising 47 HC and 16 index cases who were each an index or co-primary case to a bubble class. The number of households exceeded the number of bubbles that participated because, in four settings, HC agreed to take part, but the relevant schools withdrew. In one setting, the school agreed to take part, but the HC withdrew; a separate case from the same class was identified by community testing however, and their HC were recruited. For setting E, HC of two of the three newly identified SC infections were included. All of the index cases were symptomatic except these two.

Of the HC, 3 children and 9 adults were already reported to be infected at the start of sampling. Initial analysis focussed on HC who were considered naïve (n=35) i.e. were not reported to be infected at the start of sampling, of which 11/35 were children. (Table 3)

**Table 3.**
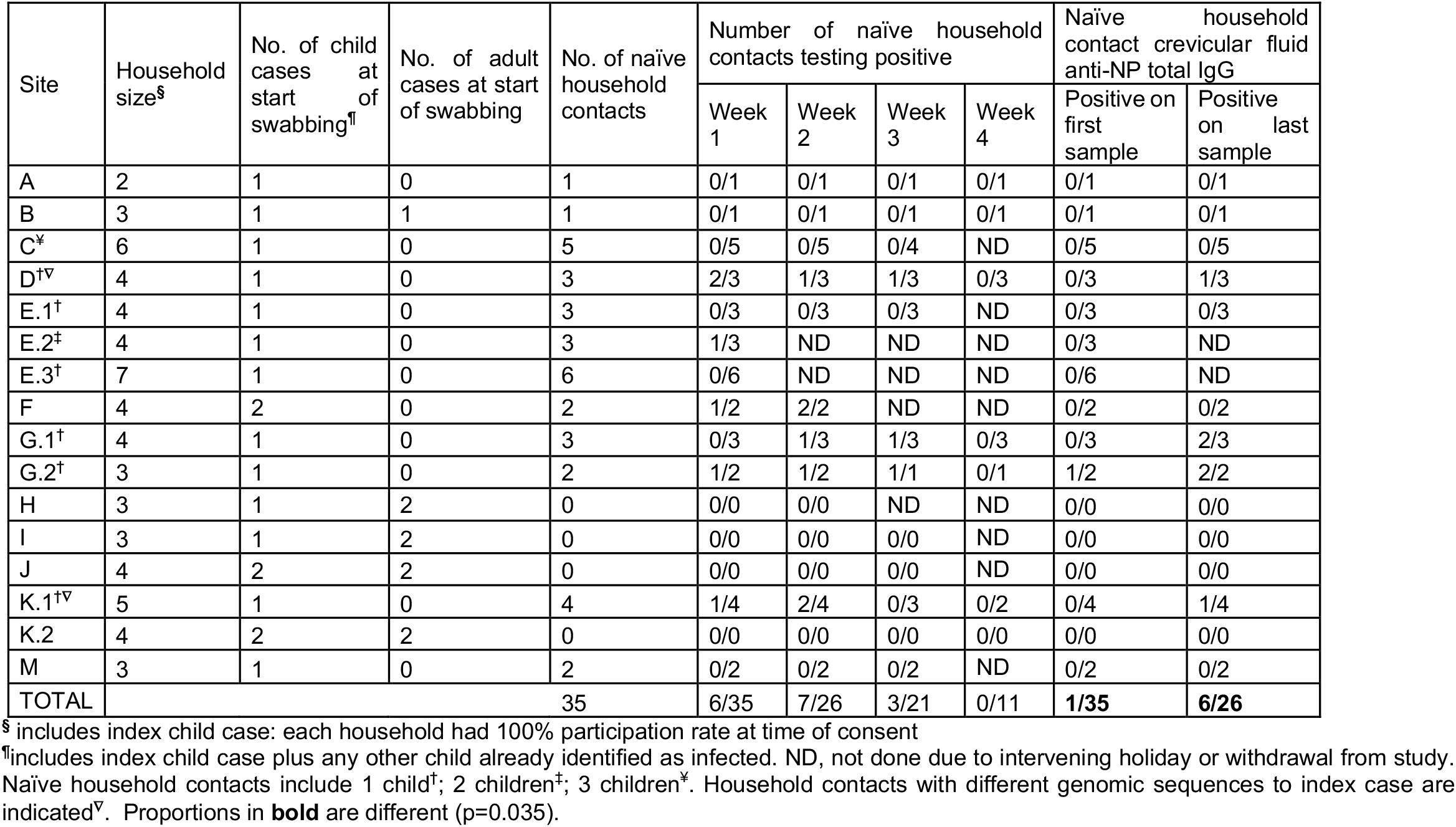
Transmission events in participating household contacts in each setting.

Over the sampling period, 9 new infections were detected among naïve HC in 8 adults and 1 child (Table 3, Figure 1C). In two households, genome sequencing revealed that the index case was unrelated to the new adult HC infections (2 per household), hence these represented secondary introduction from the community (Table 3, Appendix p5). In all other households genome sequencing was consistent with clonal household transmission (Appendix p5). Transmission by children therefore resulted in infection of 5/35 (14.3%) naïve HC. Only 1/35 (2.9%) GCF samples suggested prior COVID-19 exposure among naïve HC at the start of sampling though this rose to 6/26 (23.1%) by the end of sampling (p=0.035). Just 6 HC had been partially or fully-vaccinated; these were 2 adults each in settings K1, K2, and M.

Twelve HC who were reported to be already-infected prior to study team arrival were also sampled sequentially, but were not included in the initial analysis, due to uncertainty of transmission direction. To gain greater insight into the frequency of secondary attack rate, symptom and testing history were reviewed. Three child HC were reported to be positive prior to research sampling; based on symptom onset and date of testing, it was deduced that these child HC had been secondarily infected by the index pupil in the home. Nine adults (from 5 households) were reported to be positive prior to research sampling. For 5/9 adults, test results and/or symptoms pre-dated that of the index child, suggesting that the child was not the index case within the household. For 4/9 adults, their infection was believed to arise from the index child. Taking these additional cases into consideration, the 16 index children resulted in 9 new cases in 47 household contacts (19.1% secondary attack rate).

### Environmental samples in schools

Surface sampling identified SARS-CoV-2 in only 4/189 (2.1%) samples from bubble classrooms; 2/127 (1.6%) samples in school contact classrooms, and 5/130 (3.8%) samples from school bathrooms. (Figure 2A-C). Where detected, viral copy numbers were at the lower limits of detection except the edge of an index child’s chair in a bubble classroom that had >10^4^ E gene copies per swab in week 1, prior to deep cleaning. The same items were sampled in each location on a weekly basis (Appendix p2-3); no item became positive on subsequent sampling. Air sampling was undertaken weekly in bubble classrooms, control classrooms, and washrooms, as soon as possible after children vacated those rooms, except when availability of equipment components limited this. Only 1/68 (1.5%) air samples was positive: This was at the limit of detection, in week 2 in a school that had experienced a number of staff infections, but in a control SC classroom not known to have any pupil COVID-19 cases.

**Figure 2.**
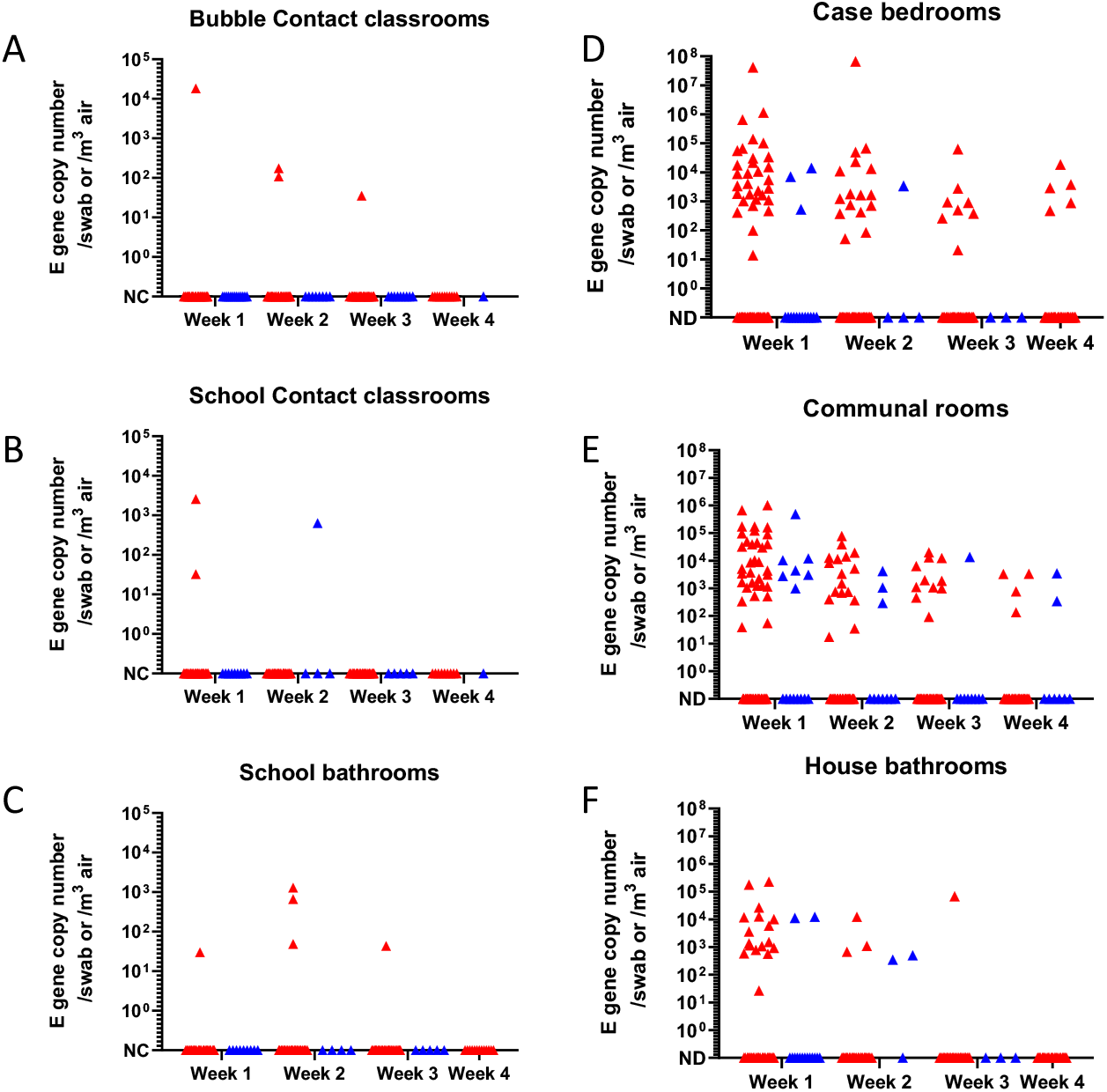
Environmental contamination with SARS-CoV-2 in schools and households. A-C, schools; D-F, households. Samples obtained at start of sampling and thereafter weekly are shown. Red indicates surface samples; blue indicates air samples. Surface and air samples were obtained from the same items and locations weekly in each school and households. Data shown as absolute E gene copy number and represent samples from 8 schools (1 SEND; 2 secondary; 5 primary) and 16 households; note y axis range differs between schools and households A. Bubble contact classroom. B. School contact classroom C. School bathroom used by bubble. D. Child’s bedroom. E. Communal room F. Bathroom used by child.

We considered the possibility that air samples might only be positive when a room is in active use. To provide context, we undertook environmental sampling in a university building (appendix p4). We identified SARS-CoV-2 in 3/10 surface samples from a small office 4 days after use by a confirmed case of COVID-19, but not in any other office or location in the same building, or on follow up (0/96 samples). We also detected low levels of SARS-CoV-2 in an air sample from the same office 4 days after use; all air samples were negative when re-tested two weeks later (Appendix p4).

### Environmental samples in households

In contrast to findings in schools, overall 262/1174 (22.3%) surface samples were found to be contaminated with SARS-CoV-2 in 16 households. Focussing on samples taken on the first visit and thereafter weekly, there was a trend to declining virus detection over time (Figure 2 D-F). The most frequent surface contamination was identified in index case bedrooms, where 60/248 (24.2%) samples tested positive, and communal rooms, where 66/241 (27.4%) samples tested positive. In bathrooms, 21/188 (11.2%) surface samples tested positive, consistent with increased bathroom surface cleaning. Personal items relating to the child such as pillows, and digital equipment such as mobile phones, remote controls and digital toys were more persistently positive over the sampling period whereas other sample types became negative within 2-3 weeks, including pet fur (Appendix p6). Surface human RNA levels were higher in households than schools (Appendix p7).

Overall, 42/150 (28%) air samples obtained in households were contaminated with SARS-CoV-2. Focussing on samples taken on the first visit and thereafter weekly, air samples were positive in 4/22 (18.2%) samples taken in the index child’s bedroom; 13/42 (30.9%) samples in the communal room; and 4/21 (19%) samples in the bathroom (Figure 2D-F). Virus levels in air were highest in the room with an infected child and infected adults. The index child and household contacts were always in the communal (living) room at the time of sampling except three settings where the index child was only in the bedroom during sampling, and one setting where the child moved between rooms. There was no apparent association between the type of dwelling (apartment or house) and air contamination. Air samples in households and schools did not differ significantly with regard to human RNA (Appendix p7).

## Discussion

Conducted during a period of enhanced precautions, transmission from index pupils to bubble contacts, and to other pupils in the school who were not close contacts, was not actively detected. Although the study was small, the findings contrasted with a secondary attack rate of at least 14.3% in household contacts of the same index cases. When household contacts who had already been tested were included in our analysis, the secondary attack rate in households with a child index case was 19.1%.

One apparent transmission incident in a class that were not isolating involved three asymptomatic pupils, who could not be linked to the original index case in that school. One of these pupils had a high viral load, leading to detection of a secondary case in a household contact and, we believe, accounted for transient low viral loads detected in two other pupils. The low viral loads were similar to environmental samples and may be consistent with transient carriage on mucosae rather than early or late infection.

Environmental surface and air sampling was conducted to understand mechanisms of transmission, where transmission occurred. This showed little or no contamination in schools including surfaces touched frequently by children, providing a high level of reassurance regarding the school environment during a period of enhanced vigilance, underlined by a difference in human RNA detection between surfaces in households and schools. This contrasted with repeated identification of virus on household items frequently touched by children, and in the air around the home, particularly where the child was present. This is perhaps not surprising since the dimensions of domestic rooms are ∼4 times smaller than classrooms and provides some insight into the risks of virus acquisition in the two settings. The detailed environmental sampling identifies digital equipment and personal items as potential fomite vectors, or as metrics of infectivity. The high proportion of air samples that were positive in the home compared with school underlines the greater risks associated with smaller rooms and is a reminder that air may remain positive for some time if not well ventilated. We considered the possibility that air sampling in schools was negative because the children were not present in the room, however control human RNA was no different in the air between schools and households. Control sampling in a different educational setting demonstrated low levels of SARS-CoV-2 RNA in the air 4 days after an office was used by a staff member with COVID-19. The low or absent levels of SARS-CoV-2 RNA in the bubble classroom also provides reassurance about the potential for ongoing infection in members of the bubble who returned to school by week 2-3.

Our findings are consistent with studies undertaken in other countries that have examined transmission in the school setting; when actively sought, transmission to bubble contacts was uncommon, with 1-2% co-primary or secondary infections identified where larger numbers have been sampled (16, 17). It is also consistent with the ∼1.5% asymptomatic infection rate reported in a recent cluster-randomised trial of daily lateral flow-testing in bubble contacts (18). The infrequency of transmission to other pupils contrasts with transmission frequency of other respiratory infections in schools, including group A streptococcus and influenza (11,12); this may reflect the multifold interventions in place during the pandemic period, or it may reflect the heterogeneity of infection in COVID19 where most transmission is caused by only a minority of infections (19, 20).

Our study prospectively examined transmission from the same children to contacts in both schools and households; the secondary attack rate in households was higher than expected, and was in stark contrast to that seen in schools. Our findings are consistent with a recent study that reported a secondary attack rate of 25% in households even when the index case is a child (21), and a recent meta-analysis (22). While children may be less likely than adults to become secondary cases, the risk of generating secondary cases is no different whether the index is a child or adult (21, 23); this pattern is confirmed in other countries (22). Quarantine for household contacts, in place throughout our study, may increase exposure of household members to index cases unless mitigated by protective measures, noting household size has been associated with urban caseloads (24). It was notable that in all households with no onward transmission to naïve contacts, householders had ensured that the affected child was isolated from others, without sharing a bedroom, whilst still affording care and supervision.

For ethical reasons, we used GCF to screen for prior SARS-CoV2 exposure, which may under-estimate exposure compared with serum (14). Prevalence of seropositivity among school pupils reported by larger scale testing is similar to levels observed in pupils in our study (25). Due to timing of our study, just six of the adult contacts had been vaccinated. Though vaccination was reported to impact household SAR (26), a recent study suggests a lesser impact with more transmissible variants (27).

Our study adopted a forensic approach to contact tracing, to not miss infections that were cleared early, or those arising late due to ongoing transmission in the class group. We took combined nasal and pharyngeal swabs to increase opportunity for virus detection and used human RNAseP as a control to ensure that negative results could be trusted. Furthermore, almost all swabs were taken by the study team; a small number of contacts were permitted to take swabs themselves if supervised. Genome sequencing identified transmission events that were genuine while also refuting others, highlighting a risk of over- or under-estimating transmission rates when relying on PCR results alone.

The study was designed to investigate bubble sizes of ∼10-15, but interpretation of ‘bubble’ changed over time, and by autumn 2020 bubble sizes routinely included 30-200 primary- and secondary-aged pupils respectively (28). The study relied upon identification of index cases who had been attending school; as such, index cases in this study were almost all symptomatic, with the expectation that asymptomatic cases would be identified among contacts as a comparison group.

There are three key limitations to our study. Firstly, the study was conducted at a time of heightened and constantly changing interventions, in particular social distancing in schools and reduced class sizes. Transmission in schools may alter when interventions relax, as indicated by more recent epidemiological reports (29). Secondly, participation rates in contacts were very low, compared with participation rates of >40% in a previous contact tracing study (12). Deterrents to participation reported anecdotally were the legal requirement to notify newly-identified infections; quarantine impact on participants; study team making home visits; low risk in children; and inclusion of older pupils. Participation by school contacts was consistently higher than bubble contacts, underlining a resistance to home visits. Recruiting bubble contacts sent home to isolate was challenging, as schools use an array of methods to contact parents. The greatest barrier to participation was the recognition that newly-identified infections would result in quarantine for entire households or classes, such that participation was actively discouraged by some groups, in contrast to predicted responses at study inception. Finally, although our study benefitted from the objective starting point of positive index cases who attend school, there is a risk of bias in all studies that rely on voluntary participation, in terms of individual schools and participants. Representation from a larger number of participants would however require expansive recruitment.

Future research of this kind may provide more meaningful data if the results are unlinked to identifiable data, or any form of notification or requirement to isolate, i.e. without real-time reporting. With reduced interventions and advent of new variants, it may be prudent to evaluate schools-based transmission in such a silent study.

## Data Availability

Any data not included in the manuscript are available from the corresponding author upon reasonable request

## Acknowledgements

The study team acknowledge the support of the school leaders, support staff, and the participants who took part in the study as well as the NIHR Clinical Research Networks and Schools Network for supporting the schools in this study.

The input of teaching and parent advisory groups is also acknowledged, as is the practical support of key staff from Imperial College Healthcare NHS Trust, Imperial College London, and Public Health England, and the TraCK study collaborators. We are also grateful to Shamez Ladhani, Samreen Ijaz, John Poh, Justin Shute and members of the Oral Fluid team at PHE Colindale.

## Author contributions

Conceptualisation SS and RC; Study site supervision RC; Laboratory supervision GPT, WB, PF, SS; Study methodology SS, RC, GPT, WB, TL, MM; Data collection LR, JZ, AR, PW, CR, MC, MS, LM, AC, GZ, KB, HL, SR, CB, RJ, NA; Data visualisation JZ, LR; Data validation LR, GPT, JZ; Data analysis JZ, JE; Funding acquisition SS, WB, GPT, RC, TL, MM; Writing, original draft SS; Review and critical editing RC, GPT, WB. Final draft: all authors.

