## Supplementary Appendix for "Transmission of SARS-CoV-2 by children to contacts in schools and households: a prospective cohort and environmental sampling study in London"

| **Supplementary Tables** |  | **Page** |
| --- | --- | --- |
| Table S1 | Items swabbed in schools, households and university | **2** |
| Table S2 | Environmental sampling results from university | **4** |
| **Supplementary Figures** |  |  |
| Figure S1 | Whole genome sequencing results from clinical samples testing positive for SARS-CoV2 by setting | **5** |
| Figure S2 | Surface contamination with SARS-CoV2 by category over sampling period (households). | **6** |
| Figure S3 | Comparison of human target detection in household and school environmental samples | **7** |
| **Supplementary Methods** |  | **8** |
| **Supplementary References** |  | **11** |

**Supplementary Table 1 Items swabbed in schools and households and university**

| Household Surface samples | | School Surface samples | | University building –surface samples | |
| --- | --- | --- | --- | --- | --- |
| Case Bedroom | Bed frame | **Classrooms (BC or SC)** | Chair | **Offices** | Chair |
|  | Chair |  | Desk |  | Computer |
|  | Computer |  | Door handle |  | Desk |
|  | Desk |  | Hand sanitiser |  | Door handle |
|  | Door handle |  | Indoor toys |  | Food packaging |
|  | Electronic game |  | Light switch |  | Light switch |
|  | Laptop |  | Locker |  | Mug |
|  | Light switch |  | Outdoor toys |  | Printer |
|  | Mobile phone |  | Reading books |  | Clothing |
|  | Musical instrument |  | Soap dispenser |  | Stationery |
|  | Pillow |  | Stationery |  | Personal equipment |
|  | Plastic toys |  | Student diary |  | Surgical mask |
|  | School bag |  | Taps |  | Telephone |
|  | Soft toys |  | Window handle |  |  |
|  | Toy shelf |  | Work folder |  |  |
|  | Wardrobe handle |  | Work tray |  |  |
| Bathroom | Door handle | **Washrooms** | Door handle | **Laboratory** | Desk |
|  | Light switch |  | Soap dispenser |  | Door handle |
|  | Taps |  | Taps |  | Laboratory equipment |
|  | Toilet flush |  | Toilet flush |  | Refrigerator handle |
|  | Toilet seat |  | Toilet seat |  | Soap dispenser |
|  | Toothbrush and paste |  |  |  | Taps |
| Communal room | Card game |  |  | **Kitchen** | Countertop |
|  | Chair |  |  |  | Cupboard handle |
|  | Door handle |  |  |  | Kettle |
|  | Electronic tablet |  |  |  | Refrigerator handle |
|  | Laptop |  |  |  | Taps |
|  | Light switch |  |  |  | Water machine |
|  | Mobile phone |  |  | **Washroom** | Door handle |
|  | Musical instrument |  |  |  | Soap dispenser |
|  | Pet cage |  |  |  | Taps |
|  | Pet fur/feathers^Ψ^ |  |  |  | Toilet flush |
|  | Plastic bottle |  |  |  | Toilet seat |
|  | Refrigerator handle |  |  | **Lobby & Lifts** | Card reader |
|  | Sofa |  |  |  | Desk |
|  | Soft toys |  |  |  | Door handle |
|  | Stationery |  |  |  | Entry keypad |
|  | Table |  |  |  | Lift buttons |
|  | Taps |  |  |  | Stair handrail |
|  | TV buttons |  |  |  |  |
|  | TV remote |  |  |  |  |
|  | Wall mirror |  |  |  |  |
|  | Water jug |  |  |  |  |

^Ψ^Included 3x cat fur, 2x dog fur, 1x bird plumage. Abbreviations, BC, Bubble contact. SC, non-bubble school contact

**Supplementary Table 2 Environmental sampling results from university**

|  |  | **Surface** | **Air** |
| --- | --- | --- | --- |
| **Office A**^¶^ | Sampling 1**^‡^** | 3/10 | 1/1 |
|  | Sampling 2 | 0/10 | 0/1 |
|  | Total | 3/20 | 1/2 |
| **Office B**^§^ | Sampling 1 | 0/10 | 0/1 |
|  | Sampling 2 | 0/10 | 0/1 |
|  | Total | 0/20 | 0/2 |
| **Shared offices** | Sampling 1 | 0/10 | 0/1 |
|  | Sampling 2 | 0/10 | 0/1 |
|  | Total | 0/20 | 0/2 |
| **Laboratory** | Sampling 1 | 0/5 | 0/1 |
|  | Sampling 2 | 0/5 | 0/1 |
|  | Total | 0/10 | 0/2 |
| **Kitchen** | Sampling 1 | 0/5 | 0/1 |
|  | Sampling 2 | 0/5 | 0/1 |
|  | Total | 0/10 | 0/2 |
| **Toilets** | Sampling 1 | 0/10 | 0/2 |
|  | Sampling 2 | 0/10 | 0/2 |
|  | Total | 0/20 | 0/4 |
| **Lobby & Lifts** | Sampling 1 | 0/8 | 0/1 |
|  | Sampling 2 | 0/8 | 0/1 |
|  | Total | 0/16 | 0/2 |

Second sampling was undertaken 14-15 days after first sampling except in offices A and B

**^‡^** Values for surface samples were: 7589.1; 31199.7; and 4493.4 E gene copies/swab. Air sample was 3104 E gene copies/cubic metre.

^¶^Second sampling was 12d after first; ^§^Second sampling was 3d after first

**Supplementary Figures**

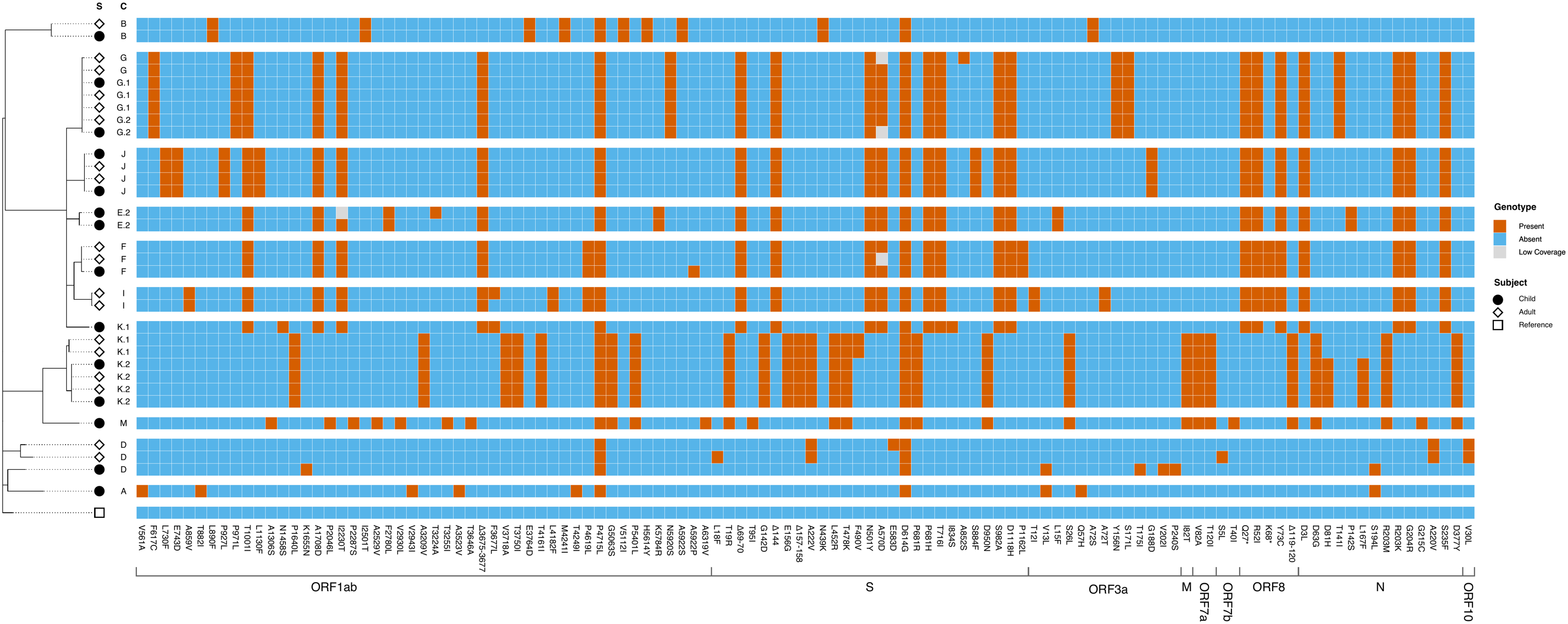
**Supplementary Figure S1** Phylogenetic relation between sequenced SARS-CoV2 isolates from participants with positive swabs

Figure S1. Phylogenetic tree and ORF mutation profile generated through whole genome sequencing of positive SARS-CoV-2 samples from TraCK study participants. The phylogenetic tree is rooted to reference sequence Wuhan-Hu-1 (GenBank accession number NC_045512.2). Samples are grouped by household cluster where possible, always considering phylogenetic tree constraints. S = Subject (Child/Adult/Reference), C = Cluster (setting or household).

**Supplementary Figure S2** Surface contamination with SARS-CoV2 in households by category over sampling period.

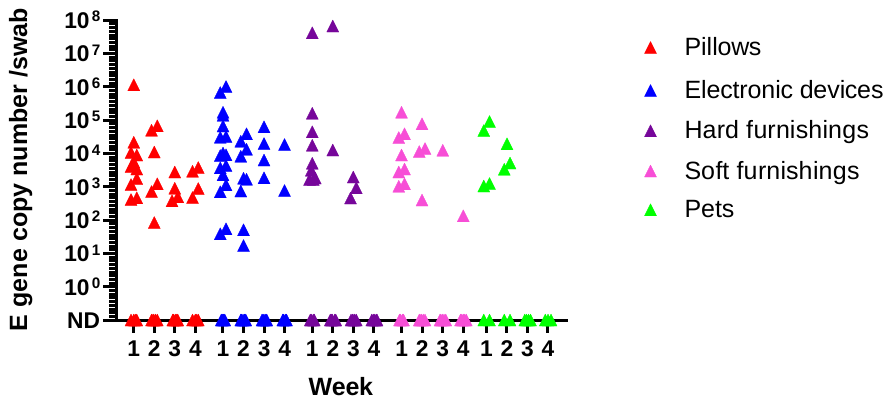

Figure S2. Environmental samples from 16 households by item category as listed in legend. E gene copy number per swab is shown for each item at each weekly time point. All items swabbed within a household were consistently sampled again on each sampling occasion within a given household; some households were swabbed for less than 4 weeks. Pet sampling included 3x cat fur, 2x dog fur, 1x bird plumage but no mucosal sampling.

**Supplementary Figure S3** Comparison of human target detection in household and school environmental samples

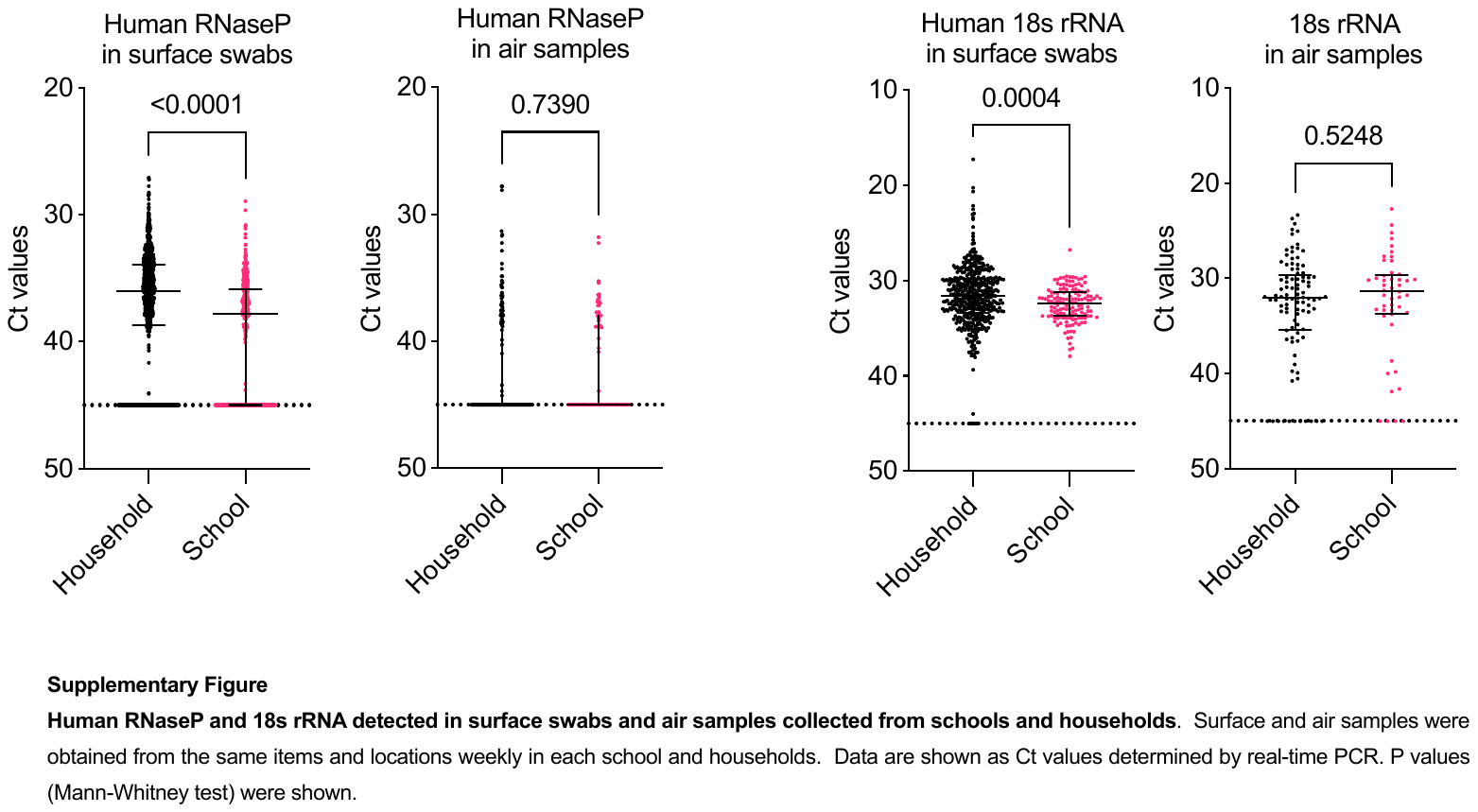

Figure S3. Human RNaseP and 18s rRNA detected in surface swabs and air samples collected from schools and households. Surface and air samples were obtained from the same items and locations weekly in each school and households. Data are shown as median and IQR Ct values determined by real-time PCR. Results between household and schools samples were compared using Mann-Whitney U test (GraphPad Prism) were shown with corresponding p values as indicated.

**Supplementary Methods**

**Context for Case and Bubble definitions and quarantine periods**

During the study period, children in England were tested for SARS-CoV-2 by PCR if exhibiting any of the recognised symptoms of COVID-19 through community or postal testing programmes. From September 1^st^ 2020 – July 19^th^ 2021, schools and nurseries were required by the UK government to undertake contact tracing for suspected or confirmed cases of COVID-19 in pupils or staff. Children with confirmed SARS-CoV-2 infection were excluded from onset of symptoms (or a positive test if no symptoms). The duration of exclusion was initially 14 days (1 Sept 2020 – 14 Dec 2020) later changing to 10 days (14 Dec – 19 July 2021). The same duration of quarantine applied to household contacts of cases regardless of vaccination status. Contacts identified by schools were excluded for the same duration. In early years and primary school settings the whole class were considered close contacts (the so-called “bubble”). In secondary school settings risk assessment identified individual close classroom contacts (face to face contact; contact within 1m for >1 minute; within 2m for >15 minutes).

**Prevailing interventions in schools and school closures**.

Schools in England re-opened in the first week of September 2020 to all children aged 5-18, having adopted a suite of preventive measures including social distancing, hand hygiene, and secondary school-aged pupils were required to wear masks when not in class; any positive cases arising in schools resulted in bubble contacts quarantining for 14 days. Schools closed in mid December 2020 for the Christmas holidays. Between January 4^th^ and March 8^th^ 2021 schools in England partially re-opened for vulnerable children, children of keyworkers, and secondary school-aged pupils undertaking exams in years 11 and 13 only. From March 9^th^ 2021 schools re-opened to all pupils and, in addition to the aforementioned measures, secondary school-aged pupils were required to undertake lateral flow antigen testing for SARS-CoV-2 twice weekly and wear masks inside and outside the classroom.

**Contact definition.**

Bubble contacts (BC) were children identified by schools who were required to isolate at home. For nurseries and primary schools, BC were in the same ‘bubble’ or class as the index case; for secondary schools, BC had been individually identified by the school as meeting the contact definitions above.

Non-bubble school contacts (SC) were children from a different ‘control’ class in the same school. SC were from a class that was adjacent in terms of age-group or geographical proximity in the school. They had not been identified by school as contacts required to isolate, but were drawn from the same wider community and, despite best efforts to keep bubbles separate, may have been exposed to similar common areas in the school as the index case the BC. Household contacts (HC) were adults and children of any age normally resident with the Case, and required to isolate.

**Sample size**.

The study was pragmatic in that it enrolled as many bubble contacts as possible within the school year. A prevalence of 25% infection was previously detected in in classroom contacts exposed to scarlet fever (1). A sample size of 40 bubble contacts was sought to detect a difference between the Null hypothesis proportion, π₀of 0.03 and the Alternative proportion, π₁, of 0.25  with 98.4% power using an exact binomial test with a nominal 5% two-sided significance level; for a sample size of 28, power was 94.49%.

**Contact sampling**

Combined nose and throat samples were obtained by the research team from each participating contact (BC, SC, or HC) as soon as possible (<48 hours) after case identification, and thereafter weekly for up to 28 days. Flocked nylon swabs (Sterilab Services, Harrogate, UK) were rubbed on the posterior fauces and then rotated gently in the nostrils no deeper than the length of the flocked end of the swab, then placed into universal transport medium. BC and HHC were sampled at home by the study team, while SC were sampled at school by the same study team. Swabs were delivered to the laboratory the same day and immediately refrigerated until processed the following working day.

**Environmental sampling***.*

For households, surface and air samples were obtained in each of three rooms (child’s bedroom, communal room, bathroom) weekly. For schools, surface and air samples were obtained from the bubble classroom, school contact classroom, and washroom weekly. Details of environmental samples obtained are listed in supplementary table 1.

For environmental surface sampling, swabs moistened in viral transport medium were used to swab 25 cm^2^ of four or five surfaces in each of three rooms (child’s bedroom, communal room, bathroom), identified as frequently touched or handled by the case, with attention on personal items (total 14 swabs). Where household pets were available, surface samples (fur or feathers) were obtained from these at the same time as other household items; mucosal samples were not obtained.

Air sampling was undertaken in the same three rooms for periods of 10 minutes (300 litres/minute, Coriolis micro, Bertin Instruments, France), with the Case present in the communal room during sampling. Environmental sampling in the home started at time of household recruitment and surfaces were re-swabbed weekly for up to 28 days at the time of household sampling.

For schools, surface swabs were taken from four or five surfaces in three locations: Bubble classroom (n=5); School contact classroom (n=5); Washroom (n=4). Schools were asked to delay cleaning of bubble classrooms until after the week 1 swabs were taken but this was not always possible. Surfaces were re-swabbed weekly for up to 28 days. Air sampling was undertaken in the same three locations, repeated weekly. Where children were present in school, sampling was undertaken immediately after children had left the class.

For the university building, surface swabs were obtained on two occasions from two academic offices; a research laboratory; washroom; kitchen area; elevator and communal lobby area.

Environmental samples were coded then tested by a research laboratory for SARS-CoV-2 RNA content using a quantitative RT-PCR detecting SARS-CoV-2 E and Orf1ab genes (2) using human RNAseP and 18s rRNA as controls for sample quality and as an indicator of human contact. Samples with high SARS CoV2 viral load (Ct value <30) were inoculated into Vero cells for culture of infectious virus as previously reported (2).

**Whole genome sequencing, lineage assignments and phylogenetic trees**

RT-qPCR was performed using an in-house protocol (3). Samples with a positive RT-qPCR result were submitted for Whole Genome Sequencing to assign lineages and generate phylogenetic trees. Samples with the highest viral loads were chosen. Automated RNA extraction was performed using a CyBio FeliX (Analytik Jena) and innuPREP Virus TS RNA Kit 2.0 (Analytik Jena) according to the manufacturer’s instructions, with a sample volume of 200 µl, without carrier RNA and with an elution volume of 50 µl. cDNA synthesis was then performed using the LunaScript RT SuperMix Kit (NEB) according to the manufacturer’s instructions with a total reaction volume of 20 µl and extracted sample volume of 5 µl. Libraries were generated using the EasySeq™ RT-PCR SARS CoV-2 (novel coronavirus) Whole Genome Sequencing kit v1 or v2 (Nimagen) according to the manufacturer’s instructions. Samples were then pooled and purified with AMPure XP (Beckman Coulter) magnetic beads. Suitable quality of libraries was confirmed using a Tapestation (Agilent) and concentrations were measured using the Qubit 1x dsDNA High Sensitivity Assay Kit (Thermofisher Scientific) and Qubit 4 Fluorometer (ThermoFisher Scientific). Pooled libraries were then diluted down to 55 pM. The final pool was then run on an iSeq 100 (Illumina) with a total of 322 cycles (151 bp paired reads and 10 bp indices). Generated fastq files were processed using the EasySeq variant pipeline (v0.6.0)(4) which is a Nextflow (5) pipeline that uses fastp (6), BWA MEM (7), SAMtools (8), BCFtools (8), LoFreq (9), mosdepth (10), BEDtools (11), SnpEff (12) and MultiQC (13) to QC, trim and assemble the reads (using reference sequence NC_045512.2) and then generate a consensus sequence and variant report before assigning a PANGO lineage (14) using pangolin (v3.1.16, lineages version 2021-10-18) (15). Sequences were aligned using Clustal Omega (16) and the alignment was then used to generate a phylogenetic tree using IQ-TREE (v2.1.3) (17). The phylogenetic tree and heatmap were generated using R (18) and the ggtree package (19).

**Gingival Crevicular Fluid** (GCF). GCF was collected from each participant at each swabbing time point (Oracol swabs, Malvern Medical, Worcester, UK). Foam swabs were rubbed on the gums for one minute at each sampling time point stored at 4° C until elution in transport medium (phosphate-buffered saline (PBS), supplemented with 10% fetal calf serum, 0.2% Amphotericin B, and 0.5% gentamicin) and then stored at -20°C until analysis by the reference laboratory (20).
